# SARS-CoV-2 vaccination of convalescents boosts neutralization capacity against SARS-CoV-2 Delta and Omicron that can be predicted by anti-S antibody concentrations in serological assays

**DOI:** 10.1101/2022.01.17.22269201

**Authors:** Alina Seidel, Bernd Jahrsdörfer, Sixten Körper, Dan Albers, Pascal von Maltitz, Rebecca Müller, Ramin Lotfi, Patrick Wuchter, Harald Klüter, Michael Schmidt, Jan Münch, Hubert Schrezenmeier

## Abstract

**Background:** Recent data on immune evasion of new SARS-CoV-2 variants raise concerns about antibody-based COVID-19 therapies. Therefore in this study the *in-vitro* neutralization capacity against SARS-CoV-2 variants Wuhan D614G, Delta and Omicron in sera of convalescent individuals with and without boost by vaccination was assessed.

**Methods and Findings:** This *in-vitro* study included 66 individuals with a history of SARS-CoV-2 infection, divided into subgroups without (n=29) and with SARS-CoV-2 vaccination (n=37). We measured SARS-CoV-2 antibody concentrations by serological assays (anti-SARS-CoV-2-QuantiVac-ELISA (IgG) and Elecsys Anti-SARS-CoV-2 S) and neutralizing titers against Wuhan D614G, Delta and Omicron in a pseudovirus neutralization assay.

Sera of the majority of unvaccinated convalescents did not effectively neutralize Delta and Omicron (4/29, 13.8% and 19/29, 65.5%, resp.). Neutralizing titers against Wuhan D614G, Delta and Omicron were significantly higher in vaccinated compared to unvaccinated convalescents (p<0.0001) with 11.1, 15.3 and 60-fold higher geometric mean of 50%-neutralizing titers (NT50) in vaccinated compared to unvaccinated convalescents. The increase in neutralizing titers was already achieved by one vaccination dose. Neutralizing titers were highest in the first 3 months after vaccination. Concentrations of anti-S antibodies in the serological assays anti-SARS-CoV-2 QuantiVac-ELISA (IgG) and Elecsys Anti-SARS-CoV-2 S predict neutralization capacity against Wuhan D614G, Delta and Omicron. While Wuhan D614G was neutralized *in-vitro* by Bamlanivimab, Casirivimab and Imdevimab, Omicron was resistant to these monoclonal antibodies.

**Conclusions:** These findings confirm substantial immune evasion of Delta and Omicron which can be overcome by vaccination of convalescents. This informs strategies for choosing of plasma donors in COVID-19 convalescent plasma programs that shall select specifically vaccinated convalescents with very high titers of anti-S antibodies.

## Introduction

The B.1.1.529 variant was first reported to the World Health Organization from South Africa on 24 November 2021 (1) and has been classified as a variant of concern (VOC), named Omicron (1).

The role of passive immune therapy of COVID-19 by convalescent plasma (CCP) is still under investigation. Data suggest efficacy of CCP in early intervention (2-6), in particular among seronegative patients and immunosuppressed patients (7-9). A significant antibody dose response has been observed in some of the CCP trials (3;4;10;11). Omicron might escape passive immune therapy since it can evade neutralization by sera from vaccinated and convalescent individuals and by monoclonal antibodies *in-vitro* (12-16) and the risk of reinfection with Omicron is higher compared to other VOC (12).

In this study, we assessed the neutralization capacity of sera from convalescents, some but not all of which were vaccinated, against the Wuhan D614G, Delta and Omicron variants. The question was whether superimmunized individuals, i.e. vaccinated convalescents, had cross-neutralization capacity against Omicron sufficient to be considered as plasma donors for passive immune therapy.

## Methods

Serum samples from 66 individuals with SARS-CoV-2 infection (with or without SARS-CoV-2 vaccination) were analyzed by two commercially available assays according to the instructions of the manufacturer (anti-SARS-CoV-2-QuantiVac-ELISA (IgG), Euroimmun, Lübeck, Germany and Elecsys Anti-SARS-CoV-2 A, Roche, Mannheim, Germany). Samples were collected after informed consent from convalescent plasma donors (17) and vaccinees. The studies were approved by the Ethical Committee of University of Ulm and Ethical Committee II, Heidelberg University.

### Preparation of pseudotyped particles

Production of rhabdoviral pseudotypes has been previously described (18). In brief, 293T cells (ATCC no. CRL-3216) were transfected with expression plasmids encoding SARS-CoV-2 spike variants Wuhan D614G (https://doi.org/10.1016/j.cell.2021.03.036), B.1.617.2 (https://doi.org/10.1126/science.abh1766), or B.1.1.529 (https://doi.org/10.1101/2021.12.12.472286) (kindly provided by Stefan Pöhlmann, Infection Biology Unit, German Primate Center, Göttingen, Germany) by Transit LT-1 (Mirus). One day after transfection, cells were inoculated with a replication-deficient vesicular stomatitis virus (VSV) vector in which the genetic information for its native glycoprotein (VSV-G) is replaced by genes encoding enhanced green fluorescent protein and firefly luciferase (FLuc) (kindly provided by Gert Zimmer, Institute of Virology and Immunology, Mittelhäusern, Switzerland), and incubated for 2 h at 37°C. Then the inoculum was removed, cells were washed with PBS and fresh medium containing anti-VSV-G antibody (I1-hybridoma cells; ATCC no. CRL-2700) added to block remaining VSV-G carrying particles. After 16-18 h, supernatants were collected and centrifuged (2,000 x g, 10 min, room temperature) to clear cellular debris. Samples were then aliquoted and stored at -80°C.

### Pseudovirus Neutralization Assay

The pseudovirus neutralization experiments were performed as previously described (18) In brief, Vero E6 cells were seeded in 96-well plates one day prior (6,000 cells/well, 2.5% FCS. Heat-inactivated (56°C, 30 min) sera were serially titrated (4-fold titration series with 7 steps + buffer only control) in PBS, pseudovirus stocks added (1:1, v/v) and the mixtures incubated for 30 min at 37°C before being added to cells in duplicates (final on-cell dilution of sera: 20, 80, 320, 1,280, 5,120, 20,480, 81,920-fold). After an incubation period of 16-18 h, transduction efficiency was analyzed. For this, the supernatant was removed, and cells were lysed by incubation with Cell Culture Lysis Reagent (Promega) at room temperature. Lysates were then transferred into white 96-well plates and luciferase activity was measured using a commercially available substrate (Luciferase Assay System, Promega) and a plate luminometer (Orion II Microplate Luminometer, Berthold). For analysis of raw values (RLU/s), background signal of untreated cells was subtracted and values normalized to cells inoculated with pseudovirus preincubated with PBS only. Results are given as serum dilution on cell resulting in 50% pseudovirus neutralization (NT50), calculated by nonlinear regression ([Inhibitor] vs. normalized response – Variable slope) in GraphPad Prism Version 9.1.1. For quantitative analyses, NT50 values <20 were set to a value of 10.

The other statistical analyses were performed using GraphPad Prism Version 9.0.2, GraphPad Software, San Diego, California USA, www.graphpad.comand NCSS 2021 Statistical Software (2021). NCSS, LLC. Kaysville, Utah, USA, ncss.com/software/ncss.

## Results

We studied a cohort of 66 individuals with a history of a SARS-CoV-2 infection (Table 1). The cohort has been subdivided in a group without vaccination (n=29) and a group with vaccination (n=37).

**Table 1:** Characteristics of the study cohort of convalescent individuals.

|  | **Prior history of infection**<br>**no vaccination**<br>**(n=29)** | **Prior history of**<br>**infection**<br>**+ vaccination**<br>**(n=37)\*** |
| --- | --- | --- |
| Median age, years (IQR) | 48 (34-57) | 48.5 (32.8-56) |
| Gender, no<br>Female / male | 13 / 16 | 26 / 11 |
| Median interval since infection, days (IQR) | 119.0 ( 85.0– 136.5) | 387.0 (226.0 – 434.0) |
| No. of vaccination doses, n (%) |  |  |
| 0 | 29 (100%) | - |
| 1 | - | 17 (45.9%) |
| 2 | - | 20 (54.1%) |
| Type of vaccination, n (%) | n.a. |  |
| BNT162b |  | 25 (67.6) |
| mRNA-1273 |  | 7 (18.9) |
| ChAdOx1 |  | 5 (13.5) |
| Median interval since last vaccination | n.a. | 93.0 (25.0 – 160.3) |

n.a.: not applicable
\* n=3 individuals had breakthrough infection after vaccination.

Non-vaccinated individuals with a history of infection exhibited Wuhan D614G neutralizing titers of 249 (geometric mean neutralizing (GMN) titers, 95% confidence interval (95%-CI) 162-381) (Fig.1A). Neutralization of Delta and Omicron was undetectable (i.e. below a titer of 20) in 4/29 (13.8%) and 19/29 (65.5%) of convalescent individuals with GMN titers of 55 (35-88) and 20 (13-32)(p=0.0021 compared to Wuhan D614G). However, convalescents who had received at least one vaccination dose exhibited significantly higher neutralizing titers compared to non-vaccinated convalescents: 2761 (1763-4323) against Wuhan D614G, 841 (545-1300) against Delta and 1204 (634-2284) against Omicron (Fig.1A). Fold-difference in GMN titers of vaccinated convalescents versus non-vaccinated convalescents is 11.1-fold for Wuhan D614G, 15.3-fold for Delta and 60-fold for Omicron (Fig.1A). Neutralizing titers against Wuhan D614G and Omicron did no longer differ significantly in convalescents after vaccination (Fig.1A). For all three variants, neutralizing titers were not significantly different between subjects who received either one or two vaccinations, resp. (Fig.1B). Neutralizing titers against Wuhan D614G and Delta were significantly higher among those with an interval ≤ 90 days since the last vaccination dose as compared to intervals >90 days, while for Omicron no such significant difference was found (Fig.1C). The geometric means of NT50 against Wuhan D614G were 4741 (≤90 days) and 1644 (>90 days)(p=0.0326), against Delta 1496 (≤90 days)) and 501 (>90 days)(p=0.0202), and against Omicron 1953 (≤90 days)) and 843 (>90 days)(p=0.1352). While in the pseudovirus neutralization assay Omicron was neutralized *in-vitro* by the polyvalent antibodies of convalescent, vaccinated individuals (Fig.1A), it was resistant to the monoclonal antibodies Bamlanivimab, Casirivimab and Imdevimab (Fig.1D).

**Figure 1:**
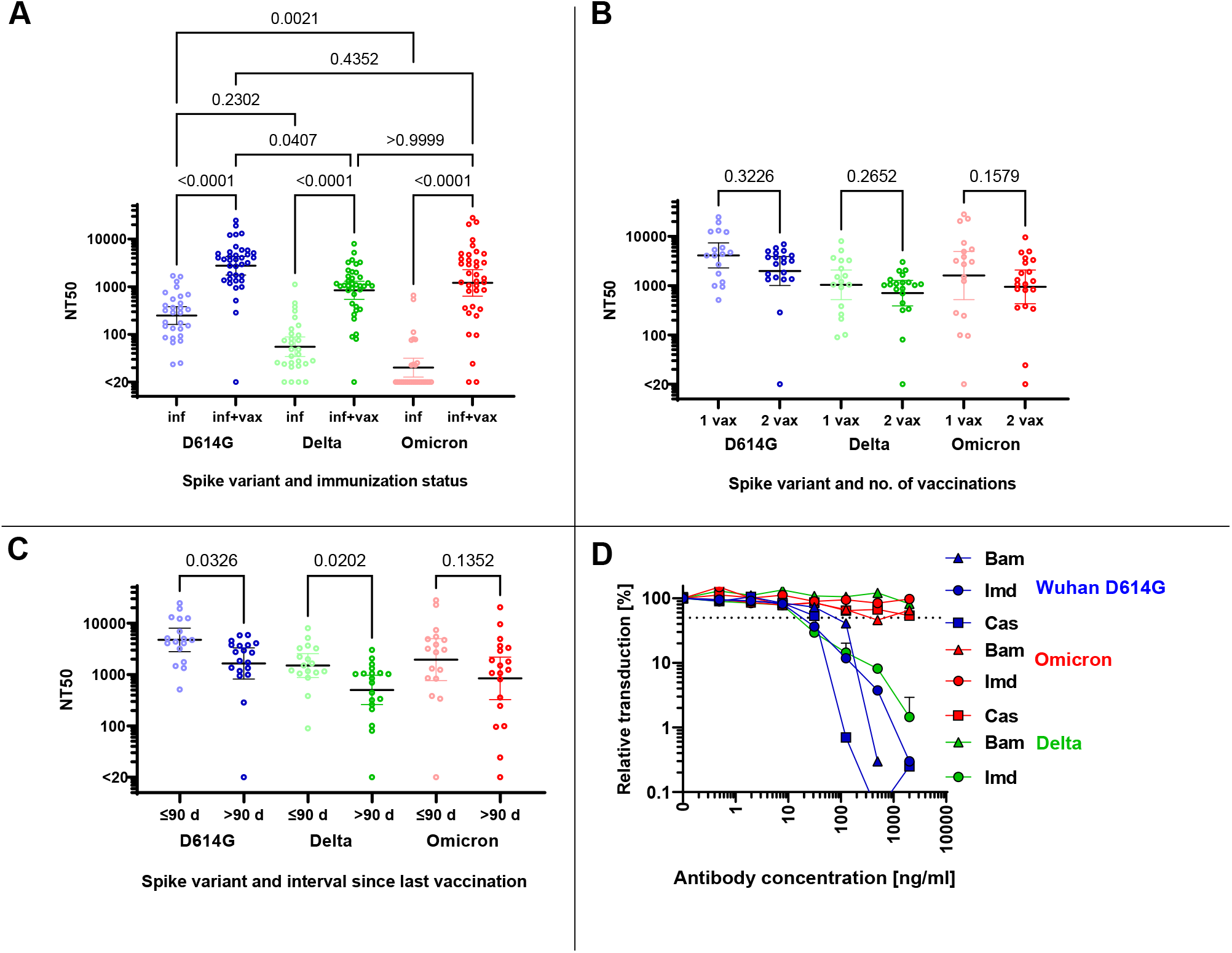
Neutralization of Spike variants by convalescent sera and monoclonal antibodies. A) NT50 for in-vitro neutralization of Wuhan D614G (blue symbols), Delta (green symbols) and Omicron (red symbols) for individuals with a history of infection (inf)(lighter colors)(n=29) and history of infection and vaccination (inf + vax, darker colors)(n=37). The p-values for the pairwise comparisons shown above the graph were calculated by Kruskal-Wallis Test followed by Dunn’s test for correction for multiple comparisons. The horizontal black lines denote the geometric mean of NT50 and the error bars the 95%-confidence interval of the geometric mean. Fold-increase in geometric mean NT50 of vaccinated convalescents versus non-vaccinated convalescents is 11.1-fold for Wuhan D614G, 15.3-fold for Delta and 60-fold for Omicron. B) NT50 for in-vitro neutralization of Wuhan D614G (blue symbols), Delta (green symbols) and Omicron (red symbols) for vaccinated, convalescent donors stratified by number of vaccination doses: 1 vaccination dose (1 vax)(lighter colors)(n=17) and 2 vaccination doses (2 vax)(darker colors)(n=20). The p-values for the pairwise comparisons were calculated by Kruskal-Wallis Test. The horizontal black lines denote the geometric mean of NT50 and the error bars the 95%-confidence interval of the geometric mean. The geometric means of NT50 were as follows: against Wuhan D614G 4102 (1 vax) and 1972 (2 vax), against Delta 1038 (1 vax) and .704 (2 vax), against Omicron 160 (1 vax) and 943 (2 vax). C) NT50 for in-vitro neutralization of Wuhan D614G (blue symbols), Delta (green symbols) and Omicron (red symbols) for vaccinated, convalescent donors stratified by interval between vaccination and collection of serum sample: ≤ 90 days (lighter colors)(n=17) and >90 days (darker colors)(n=19). The p-values for the pairwise comparisons were calculated by Kruskal-Wallis Test. The horizontal black lines denote the geometric mean of NT50 and the error bars the 95%-confidence interval of the geometric mean. The geometric means of NT50 were as follows: against Wuhan D614G 4741 (≤90 days) and 1644 (>90 days), against Delta 1496 (≤90 days)) and 501 (>90 days), against Omicron 1953 (≤90 days)) and 843 (>90 days). D) Inhibition of cell entry of Wuhan D614G (blue symbols), Delta (green symbols) and Omicron (red symbols) spike carrying pseudoparticles by monoclonal antibodies. Increasing doses of Bamlanivimab (triangles), Casirivimab (squares) and Imdevimab (circles) were preincubated with pseudoparticles before addition to cells (doses were titrated in 4-fold dilution from 2000 ng/ml to 0.49 ng/ml (final concentrations on cells)). Infection rates in A-D were determined 16 hours post infection by measuring luciferase activities in cellular lysates. Data shown were derived from one experiment performed in duplicates.

The correlation matrix of NT50 against Wuhan D614G, Delta and Omicron, the –QuantiVac-ELISA (IgG) ELISA and the Elecsys Anti-SARS-CoV-2 S revealed good correlations between all five assays, in particular between the two anti-SARS-CoV-2 serological assays (Spearman correlation 0.95) and the NT50 against Wuhan D614G and Delta (Spearman correlation 0.93)(Fig.2). Spearman correlation between anti-SARS-CoV-2-QuantiVac-ELISA (IgG) and NT50 against Omicron or Delta was 0.88 and 0.94 (Fig.3A and Fig.3C). The Spearman correlation between Elecsys Anti-SARS-CoV-2 S and NT50 against Omicron and Delta was 0.92 and 0.90 (Fig.3B and Fig.3D).

**Figure 2:**
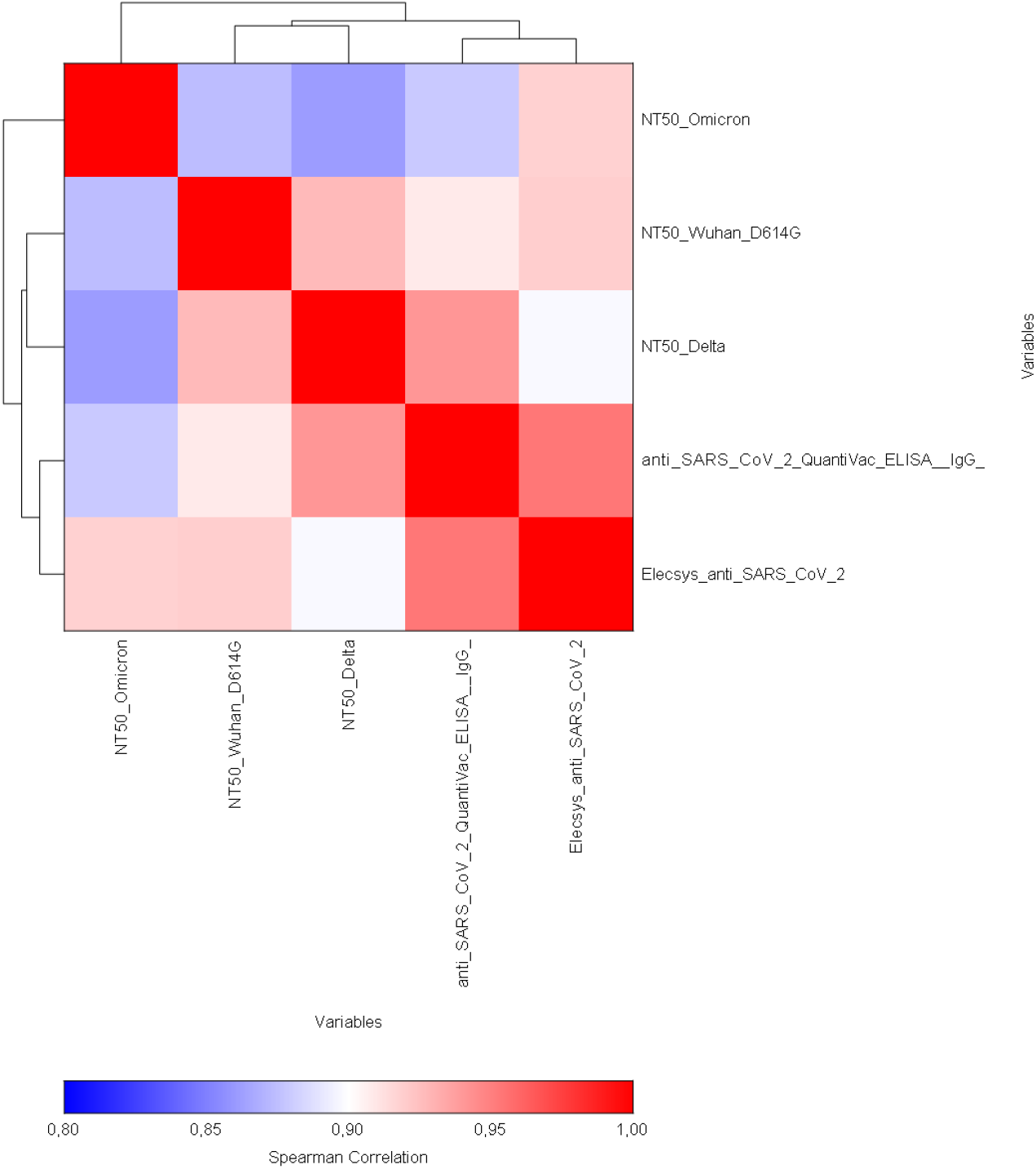
Correlation of anti-S antibody concentrations and neutralization capacity against spike variants. Correlation matrix of NT50 against Omicron, Wuhan D614G, Delta, and anti-SARS-CoV-2 QuantiVac (IgG) ELISA and Elecsys SARS-CoV-2 based on Spearman Correlation. The dendrogram shows clustering of NT50 against Wuhan D614G and Delta and between anti-SARS-CoV-2-QuantiVac-ELISA (IgG) and Elecsys Anti-SARS-CoV-2 S.

**Figure 3:**
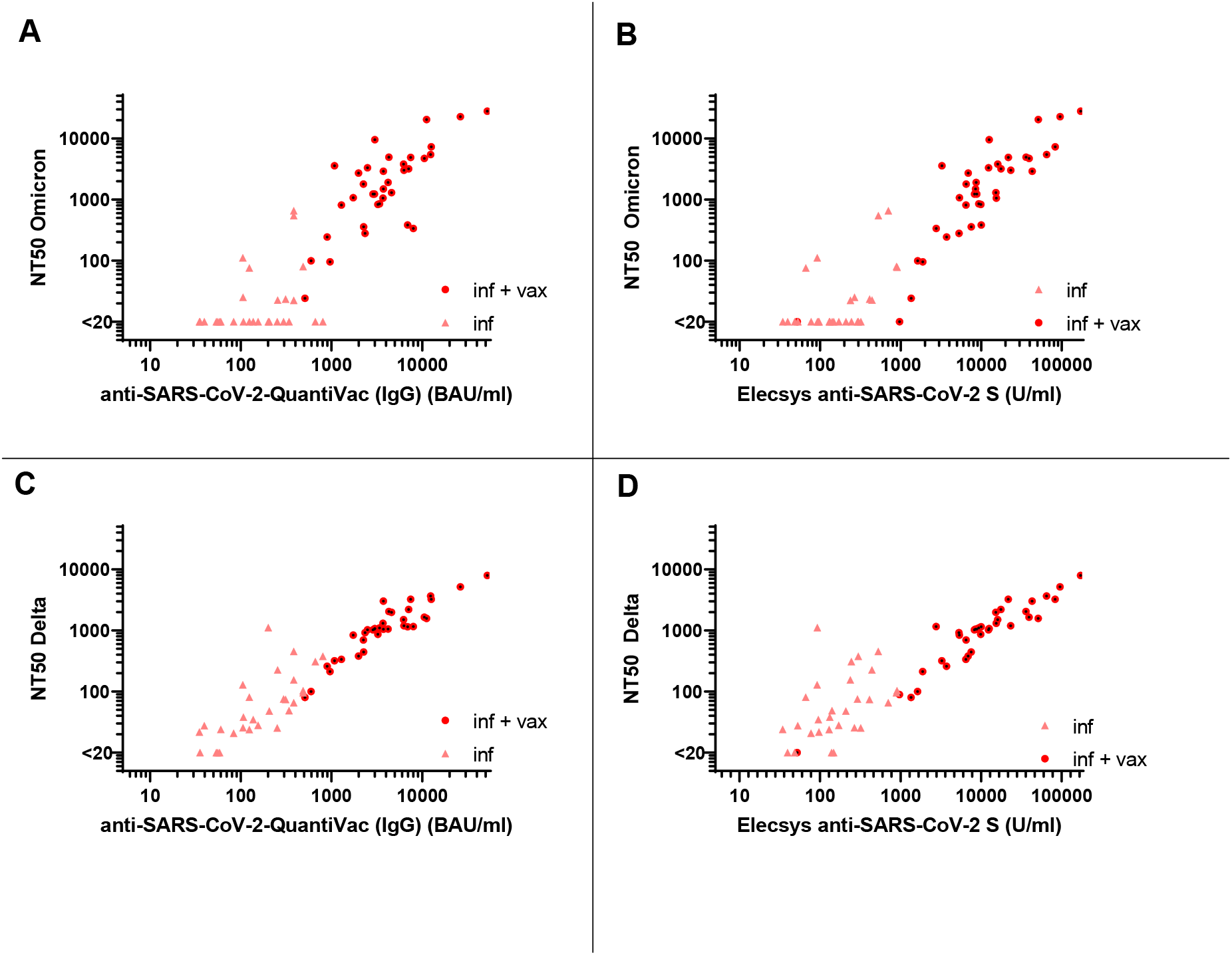
Correlation between anti-S antibody concentrations and NT50 against Delta and Omicron. A) Correlation between anti-S antibody concentrations measured by anti-SARS-CoV-2-QuantiVac-ELISA (IgG) and NT50 against Omicron (Spearman correlation 0.88). Results of non-vaccinated convalescents (inf) are shown as triangles, and results of vaccinated convalescents (inf + vax) are shown as filled circles. B) Correlation between anti-S antibody concentrations measured by Elecsys Anti-SARS-CoV-2 S and NT50 against Omicron (Spearman correlation 0.92). Results of non-vaccinated convalescents (inf) are shown as triangles, and results of vaccinated convalescents (inf + vax) are shown as filled circles. C) Correlation between anti-S antibody concentrations measured by anti-SARS-CoV-2 QuantiVac-ELISA (IgG) and NT50 against Delta (Spearman correlation 0.94). Results of non-vaccinated convalescents (inf) are shown as triangles, and results of vaccinated convalescents (inf + vax) are shown as filled circles. D) Correlation between anti-S antibody concentrations measured by Elecsys Anti-SARS-CoV-2 S and NT50 against Delta (Spearman correlation 0.90). Results of non-vaccinated convalescents (inf) are shown as triangles, and results of vaccinated convalescents (inf + vax) are shown as filled circles.

Plasma units for immune therapy shall have very high neutralizing titers -we assumed NT50≥640. Receiver operating characteristics (ROC) demonstrate that both anti-SARS-CoV-2-QuantiVac-ELISA-(IgG) and Elecsys anti-SARS-CoV-2 S excellently predict these neutralizing titers with areas under the curve between 0.94 and 0.95 for anti-SARS-CoV-2-QuantiVac-ELISA (IgG) and between 0.95 and 0.98 for Elecsys Anti-SARS-CoV-2 S (Fig.4A-C).

**Figure 4:**
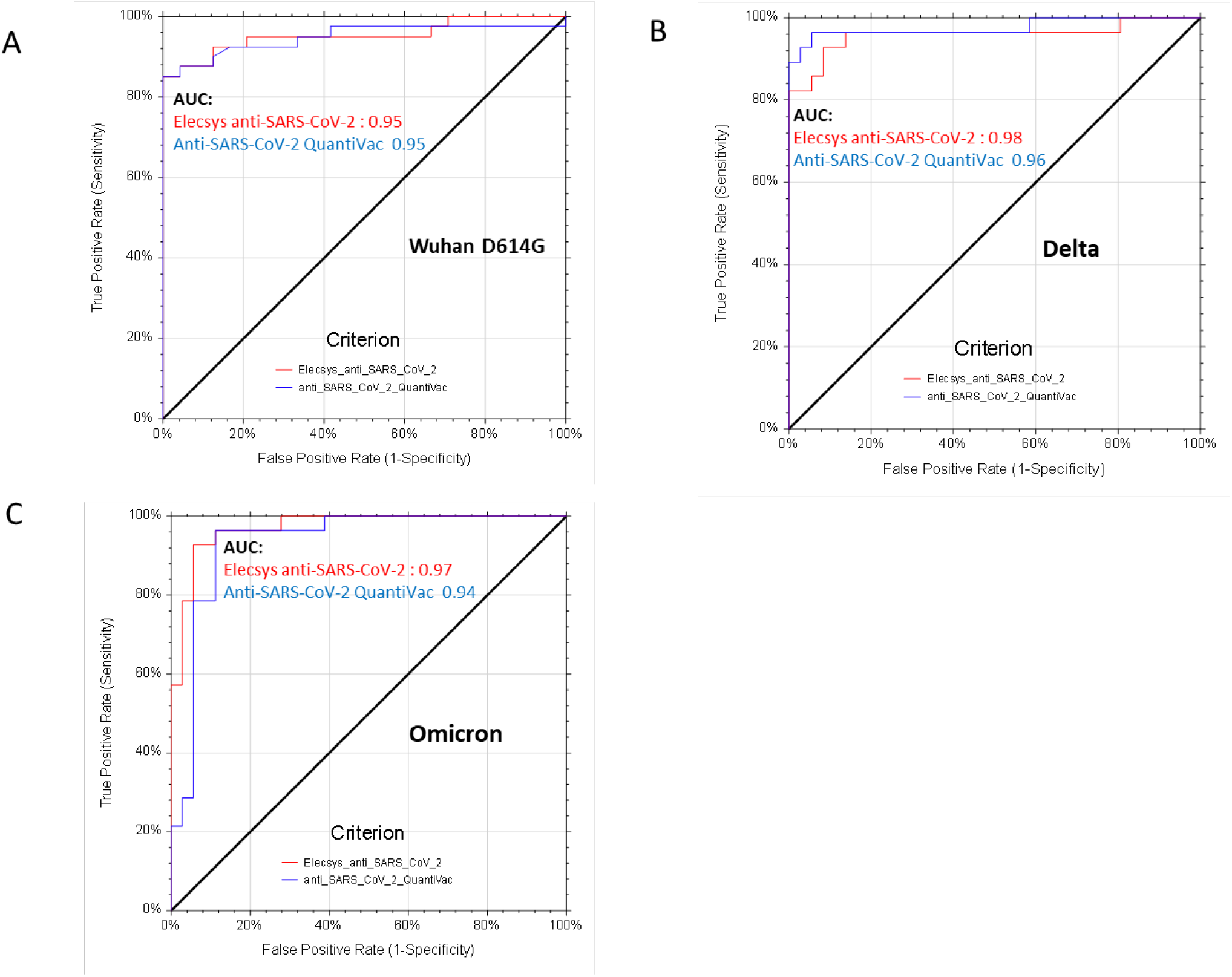
Receiver operating characteristics analyses of serological assays and neutralization of spike variants. Receiver operating characteristics (ROC) analyses of Elecsys Anti-SARS-CoV-2 S (red lines) and anti-SARS-CoV-2-QuantiVac-ELISA (IgG) (blue lines) prediction of neutralization of Wuhan D614G (panel A), Delta (panel B) and Omicron (panel C). A positive neutralizing titer was arbitrarily defined as ≥640. Area under the curve (AUC) is reported in the graphs, p<0.0001 for all serological assays and all spike variants.

## Discussion

Our observations are consistent with other *in-vitro* studies which reported significant immune evasion by Omicron (12;13;15;16). This has raised concerns that antibody therapies may no longer be effective against Omicron.

Therefore, in contrast to other recent studies on Omicron immune evasion, we focus on convalescent individuals and the implication of evasion from antibody-mediated neutralization for future CCP programs. Our data confirm *in-vitro* resistance of Omicron to several monoclonal antibodies used in clinical practice (13-16) questioning the efficacy in Omicron infected patients. Also, Delta and Omicron are no longer well neutralized *in-vitro* by sera of convalescents from the first and second surge of the SARS-CoV-2 pandemic. However, in convalescents just one dose of SARS-CoV-2 vaccination can restore *in-vitro* neutralization capacity against Delta and Omicron. In contrast to other recent reports on significantly lower neutralization capacity in vaccinated convalescent donors (14-16) compared to wild type, we observed a comparable neutralization capacity against Wuhan D614G and Omicron. This might be due to our high sample size, the neutralization assay, the large proportion of donors with high anti-S antibody concentration in our cohort and the vaccination scheme. Our findings suggest that even without adaption of current available vaccines, the broader immune repertoire in superimmunized individuals can cover novel variants (19), particularly in the first three months after vaccination when the highest neutralizing titers are achieved.

Systematic screening of convalescent, vaccinated donors using commercially available high-throughput serological assays (Anti-SARS-CoV-2-QuantiVac-ELISA (IgG); Elecsys Anti-SARS-CoV-2 S) can identify plasma donors with very high SARS-CoV-2 antibody concentrations, who also have very high *in-vitro* neutralizing titers against Wuhan D614G, Delta and Omicron. Therefore, for future convalescent plasma programs, priority should be given to superimmunized donors with previous infection plus at least one dose of a SARS-CoV-2 vaccination (preferably within the last 3 months) with very high SARS-CoV-2 antibody concentrations as measured by serological assays. A concept of early, very high titer CCP from highly selected superimmunized donors must be investigated in clinical trials (e.g. COVIC-19 trial, EudraCT 021-006621-22).

## Data Availability

All relevant data are within the manuscript and its Supporting Information files.

## Acknowledgement

The clinical trials COVIC-19 (EudraCT 021-006621-22) is supported by the Bundesministerium für Bildung und Forschung (“German Federal Ministry of Education and Research”) and the randomized clinical trial CAPSID (EudraCT including the preparation of convalescent plasma units was supported by the Bundesministerium für Gesundheit (“German Federal Ministry of Health”). H.S., J.M. and H.K. acknowledge funding from the Ministry for Science, Research and the Arts of Baden-Württemberg, Germany (COVID-19 Sonderförderlinie, CORE Project). HS acknowledges funding by the European Commission (HORIZON2020 Project SUPPORT-E, no. 101015756). J.M. further acknowledges funding by EU’s Horizon 2020 research and innovation programme (Fight-nCoV, 101003555).

